# ANTIMICROBIAL SUSCEPTIBILITY PATTERNS OF OF TRACHEAL TUBE ISOLATES FROM PATIENTS OF VENTILATOR ASSOCIATED PNEUMONIA (VAP) ADMITTED TO THE ICU OF A TERTIARY CARE HOSPITAL IN RAWALPINDI PAKISTAN

**DOI:** 10.1101/2023.04.12.23288476

**Authors:** Miriam Farooq Baba, Ramal Abdullah, Soman Nadim Iqbal, Maryam Tariq, Muhammad Moaaz Ali, Mohammad Hassan, Fatima Kaleem, Shahid Ahmed Abbasi

**Affiliations:** Foundation University Medical College (FUMC), Foundation University Islamabad (FUI), Pakistan; Fauji Foundation Hospital, Foundation University Medical College FUMC, FUI, Pakistan; Pathology department, Fauji Foundation Hospital (FFH), Rawalpindi, Pakistan; Department of Pathology, Foundation University Medical College (FUMC), Foundation University Islamabad (FUI), Pakistan

## Abstract

The respiratory system is a complex interconnected functionality which consists of a series of branching tubules and the lungs containing many alveoli. The alveoli are highly vascularized to provide a large surface area for gaseous exchange. The investigation was conducted in the Fauji Foundation Hospital’s Department of Microbiology from November 2019 to June 2022.. The various bacterial strains that were recovered from a total of 802 mechanically intubated individuals make up the data presented in this study. *Acinetobacter* spp 270 (34%), *Pseudomons* spp 165(21%), and *Klebsiella* spp 95 (12%) were the most frequently isolated bacteria. Main Gram-positive organisms isolated included *Methicillin-Resistant Staphylococcus aureus* 66 (8%), Enterococcus 48 (6%). *MRSA* was 100% sensitive to Vancomycin, but only 97% sensitive to the Linezolid. The *Acinetobacter* spp found in the tracheal tubes was reported to be 77% sensitive to Polymyxin B.

## INTRODUCTION

The term ventilator associated pneumonia (VAP) describes a pulmonary parenchymal infection occurring in patients subjected to invasive ventilation for at least 48 hours. It remains to date, one of the most frequently occurring infections among patients in need of mechanical ventilation (1). Numerous host defences are continuously at work to prevent the occurrence of diseases. In the airway, they comprise the hair cells, the secretory goblet cells which secrete IgA and mucous responsible for clearance of inhaled antigens. The mucosa comprises dendritic cells, plasma cells, macrophages and T lymphocytes responsible for immune monitoring and removal of invading pathogens. In the lung parenchyma, the defence is mostly offered by the alveolar macrophages along with small amounts of B and T lymphocytes (2)

One of the major contributors to VAP development is an endotracheal tube (ETT). Microbes can enter the trachea through microaspiration or by developing a biofilm on the interior surface of the ETT.(3). Every time the ETT seal is compromised micro-aspirations occur. Factors affecting micro-aspirations include subglottic secretions, ETT cuff pressure, folds in the ETT and ETT movement. These contaminated aspirations render the patient susceptible to a number of respiratory tract problems (4). The invading microbes move along the airflow and gravity to contaminate the distal airways. Factors including impaired cough reflex, compromised muco-ciliary clearance and damaged tracheal epithelium, create favourable circumstances for bacterial invasion and formation of biofilms on the ETT surface (5)

Furthermore, the insertion of ETT often produces ijury and implanation of exogenous and endogenous bacterial inoculum in the tracheal mucosa. It provides an adhering surface to the microorganisms and serves as a reservoir forming a biofilm (6). A biofilm is an organised colony of bacteria that is connected to an inert or living surface and encapsulated in a self-produced polymeric matrix. (7The development of biofilms on the surface of ETT has now been demonstrated to be an international occurrence that is closely linked to the pathogenesis of ventilator-associated pneumonia (VAP) (8).

The concept “nosocomial pneumonia,” also known as “ventilator associated pneumonia,” or “VAP,” refers to a patient who has acquired it after 48 hours of supplemental ventilation with an endotracheal tube or tracheostomy. Patients who require intubation have a six-to twenty-fold increased risk of developing pneumonia, and their overall mortality rates are between 20 and 40% (9). Furthermore, VAP affects 5–40% of patients who have been receiving invasive mechanical ventilation for more than two days, with major variations depending on the population, kind of ICU, and diagnostic criteria utilized (1). The frequency of VAP in south-east Asian nations ranges from 2.13 to 116 per thousand ventilator days, according to a systemic review and meta-analysis of the subject (10).

Early onset (EOVAP), which occurs within 5 days of hospitalisation and has a better prognosis, and late onset (LOVAP), which manifests beyond 5 days of hospitalisation and is primarily caused by multi-drug resistance (MDR) organisms, are two categories for VAP. (11). Bacteria causing early-onset VAP include *Streptococcus pneumoniae, Haemophilus influenzae*, methicillin-sensitive *Staphylococcus aureus* (MSSA), antibiotic-sensitive enteric gram-negative bacteria (GNB) like *Escherichia coli, Klebsiella pneumoniae, Enterobacter* spp., *Proteus* spp., and *Serratia marcescens* while bacteria causing late-onset VAP include MDR bacteria such as methicillin-resistant *Staphylococcus aureus (*MRSA), *Acinetobacter baumannii* and *Pseudomonas aeruginosa* (12).

In this study we aim to establish the antimicrobial susceptibility of the resistant pathogens present in ICU endotracheal tubes for the identification of key pathogens which need our immediate attention in order to promote appropriate treatment options. The detection, prevention and management of antimicrobial resistance is very crucial for the subsistence of the effective drugs available at present in our health care system and antibiotic clinical pipeline as potential treatment options for infective diseases (13)

## MATERIALS AND METHODS

This observational study was conducted at the Fauji Foundation Hospital in Rawalpindi’s Department of Microbiology from November 2019 to December 2022. The hospital’s ethical review committee provided their ethical approval.

All samples of ETTs from patients undergoing mechanical ventilation or intubation for a period of more than 48 hours, and showing clinical findings of ventilator acquired pneumonia, in the intensive care unit the hospital were included in the study. Duplicate samples of ETT from wards other than ICU were excluded from the study along with patients who were already started on broad spectrum antibiotics.Demographic details such as age and gender were recorded for each patient along with day of arrival and day of discharge from the hospital.

Samples for microbiological culture analysis were collected from the tips of endotracheal tubes (2 to 5cm in length), in sterile containers. Standard culture techniques were applied. 0.01mL of sample was placed in distilled water and was inoculated on blood and MacConkey’s agar. Samples were incubated at 37LJ for a period of 48 hours. Samples producing growth of >10^3^ CFU/ml were considered significant. The Gram staining technique was applied for the differentiation of Gram positive from Gram Negative Bacteria. For further identification, biochemical testing was performed. This included coagulase test to verify the presence of Staphylococcus aureus and tests for Gram-negative microbes such as oxidase, bile esculin hydrolysis and API 20E and API 20NE (BioMerieux). Furthermore catalase, coagulase and deoxyribonuclease test for cefoxitin resistant MRSA detection were also applied. All the isolated pathogens were identified till the species level.

We used 0.5 McFarland, uniformly distributed on a Mueller-Hinton agar plate, to compare the turbidity of the inoculum. As directed by the manufacturer, antibiotic discs were administered. The modified Kirby-Bauer disc diffusion method was used to test the antimicrobial sensitivity of all the bacteria on Mueller Hinton Agar (MHA), Oxoid. Antimicrobials were cultured with the isolates for 18 to 24 hours at 37 °C. The utilised antimicrobial disk’s advantages are as follows: Polymyxin B (PB 300 ug), Doxycyline (DO 30 ug), Gentamicin (CN 10 ug), Gentamicin (CN 120ug) for *Enterococcus spp*, Amikacin(AK 30 ug), Imipenem (IMP 10 ug), Ciprofloxacin (CIP 5 ug), Meropenem (MEM 10 ug), Trimethoprin/Sulfamethoxazole (SXT 25 ug), Sulzone (SCF), Minocycline (MH30 ug), Chloramphenicol (C 30 ug), Vancomycin (VA 30 ug), Linezolid (LZD 30 ug), Erythromycin (E 15 ug), Ceftazidime (CAZ 30 ug) and Aztreonam (ATM 30 ug).

MIC of Vancomycin in case of Gram-positive bacteria was done using E-strips. Interpretation of zone diameter and minimal inhibitory concentration (MIC) measurements of the following drugs were according to the CLSI 2022 standards (14).

All data was entered and analyzed using Python version 3.10.7

## RESULTS

The data outlined in this study consists of the different strains of bacteria that were isolated from a total number of 802 mechanically incubated patients during the years 2019-2022. The highest number of patients presented in the year 2019 followed by 2020. A higher number of female patients (652) was observed in comparison to male patients (150).

As presented in the following tables, 7 strains of bacteria were isolated on which further tests and trials were conducted, and the most effective antibiotic then concluded. The most common bacteria isolated was *Acinetobacter spp* 270 (34%) followed by *Pseudomonas* spp *165* (21%) and *Klebsiella* spp 95 (12%). Main Gram-Positive organisms isolated were *MRSA 66(8%), Enterococcus* spp *48 (6%) and Staphylococcus aureus 30 (4%)*. Others 45 (5.61%) included *Corynebacterium 19 (2%), Enterobacter* spp *15(2%)*. The remaining isolates *Escherichia Vulneris, Serratia* spp, *Citrobacter* spp, *Stenotrophomonas maltophillia were 2*.*5%*

Upon inspecting the sensitivity patterns, Polymyxin B was found to be the most effective against *Acinetobacter* infections with a 76.7% sensitivity. This was followed by a 75.6% sensitivity to Tigecycline. *MRSA* showed a 100% sensitivity to Vancomycin and a 97 % sensitivity to Linezolid.

**Figure 1:**
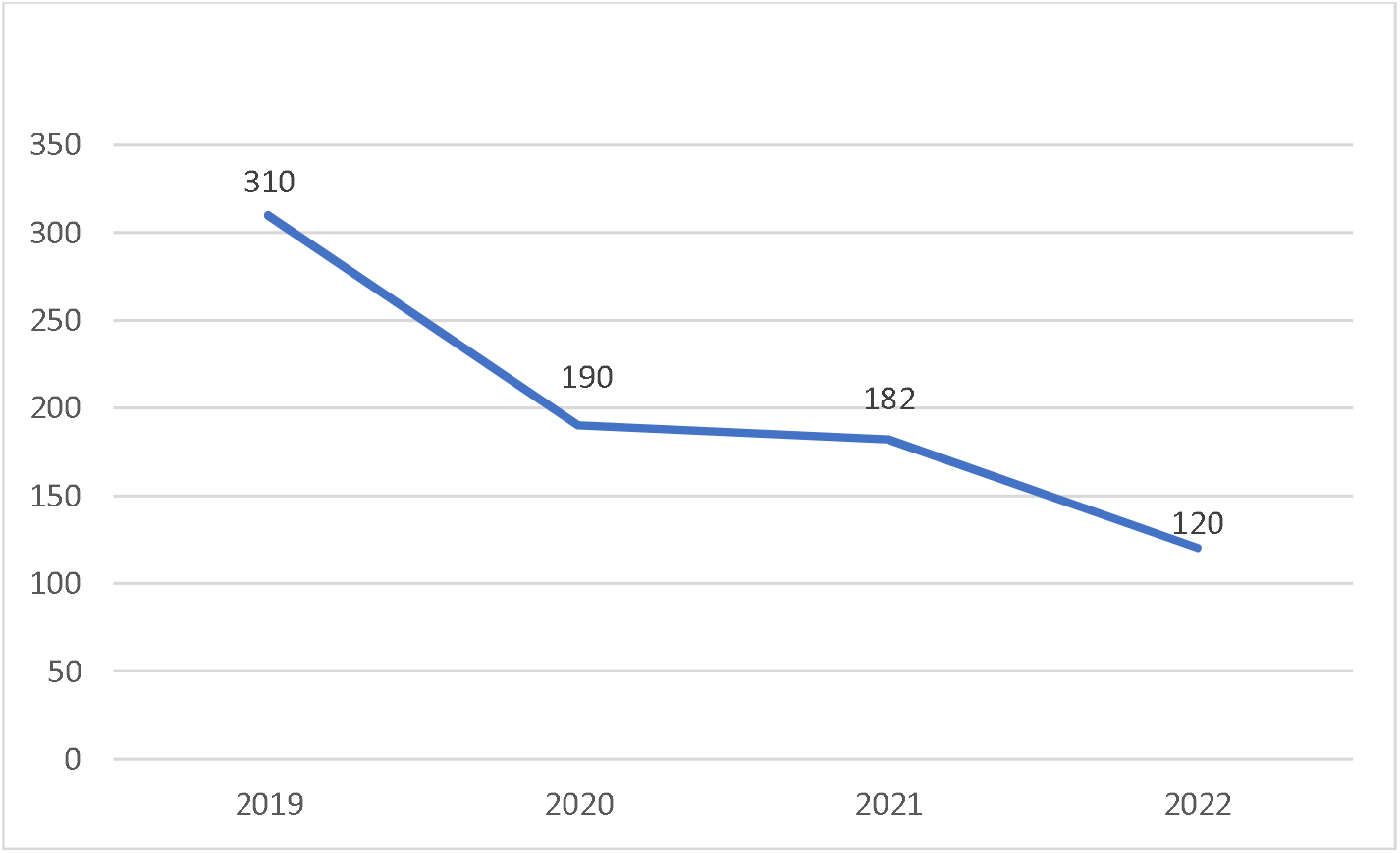
Number of Patients.

**Figure 2:**
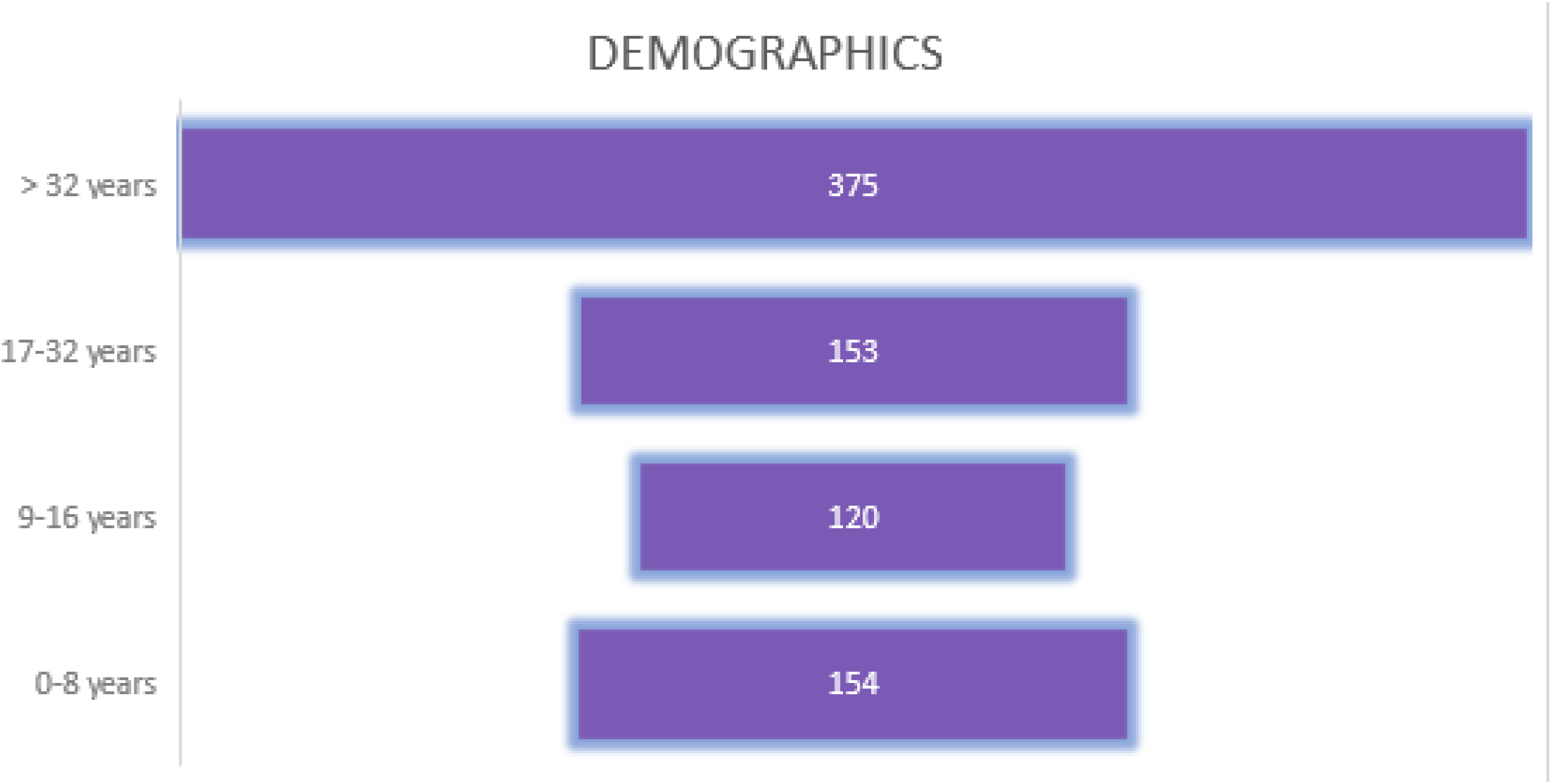
Basic Demographic Age Groups.

**Figure 3:**
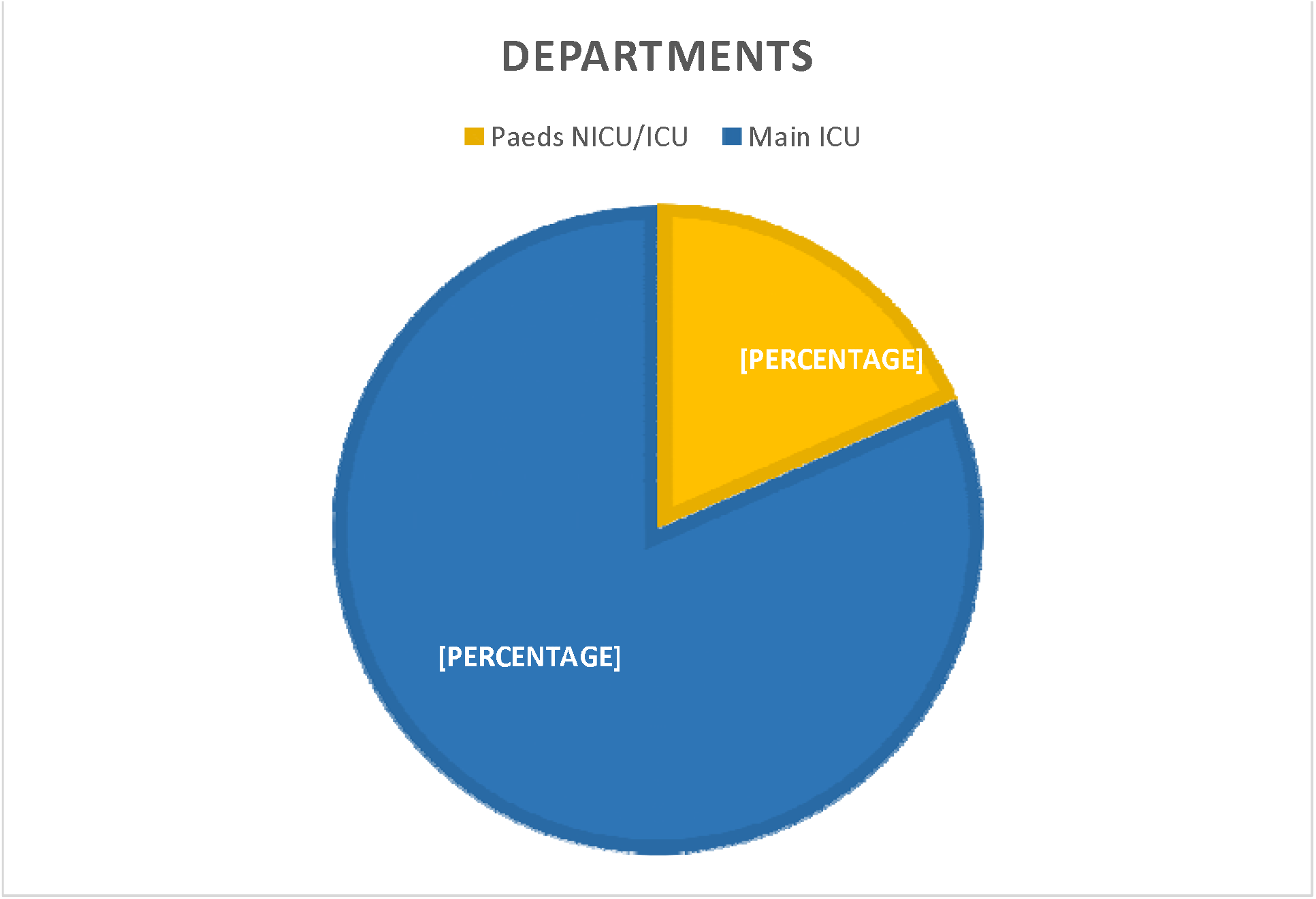
Departments.

**Figure 4.**
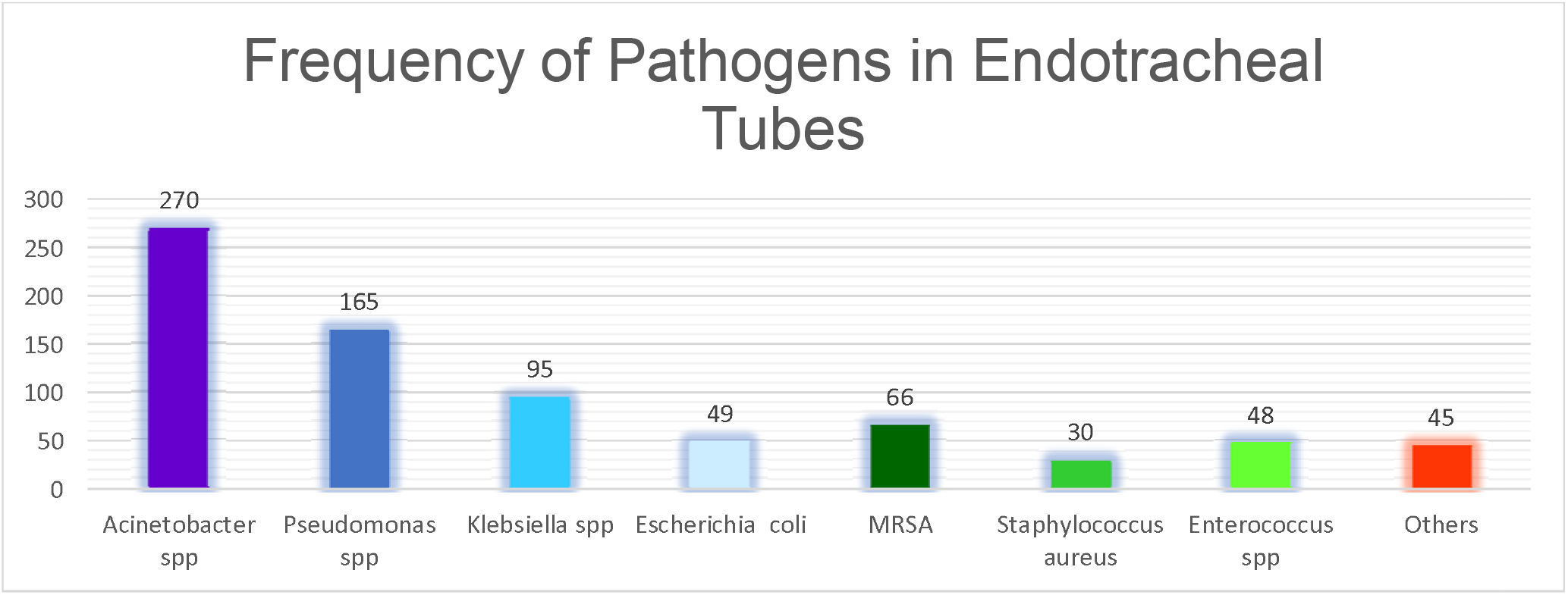
Frequency of Pathogens isolated from ETT.

## DISCUSSION

Antibiotic resistance is a global concern, as it threatens to take us back to a pre antibiotic era where developing infectious diseases will render clinicians unable to treat patients (15). According to a recent study, respiratory tract infections are the most prevalent form of clinical infections encountered in this time and a significant percentage is also common in the intensive care units (ICUs) of hospital facilities. Out of these, ventilator associated pneumonia (VAP) has a high frequency of occurrence i.e. 74.0% of the total number of pneumonia patients treated in the ICU (16). Patients of VAP are at high risk of mortality especially due to recent antibiotic resistant pathogens (17). A review of antibiotic susceptibility patterns, from tracheal tube isolates, in the ICU of our hospital may lead to a better management plan for the affected patients.

In attempts to improve upon existing studies which have been conducted in our department, a larger sample size was used and the data was collected over a longer time period of 3.5 years. VAP continues to be a concern as it remains a leading cause of death in patients coming into the ICU. The mortality rate is about 30%.

Potential reasons behind difference in data collected in 2019 compared to 2022 could boil down to shifting patterns of growth which can be attributed to many factors such as weather-based changes, SARS CoV 2 pandemic and changes in susceptibility patterns (18). *Acinetobacter* spp continues to be a concern in tertiary care as it is still endemic in hospital settings; its ever-increasing resistance to carbapenem which are the last resort antibiotics, are making it becoming harder to control (19)

Comparing the data to a similar study, the sensitivity rates have continued to decrease (20). The use of combinations of carbapenems like meropenem and imipenem with Colistin and Tigecycline would be the advisable option based on the sensitivity data in Table 1. But the adverse effects including nephrotoxicity are also a cause for concern. The sensitivity and growth patterns for some of the top species isolated from samples were looked into more detail as they account for majority of the patients in our setup. The isolates of *Acinetobacter* spp and *Pseudomonal* spp displayed a higher sensitivity to Polymyxin compared to other anti-biotics, while the *Klebsiella* spp were more sensitive to Tigecycline, Sulzone and Polymyxin.

**Table 1:**
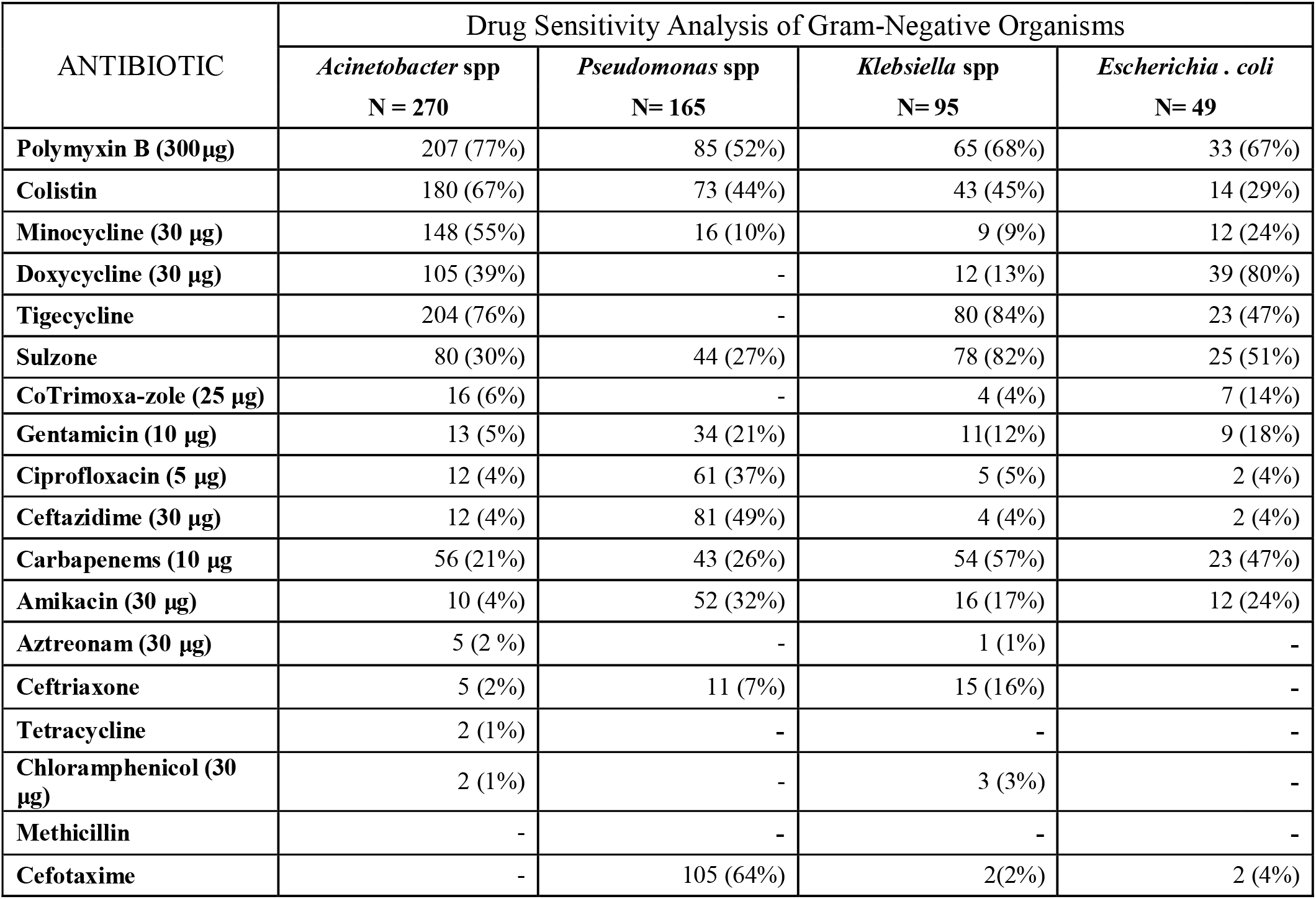
Drug Sensitivity Analysis (Gram-Negative Bacteria)

**Table 2:**
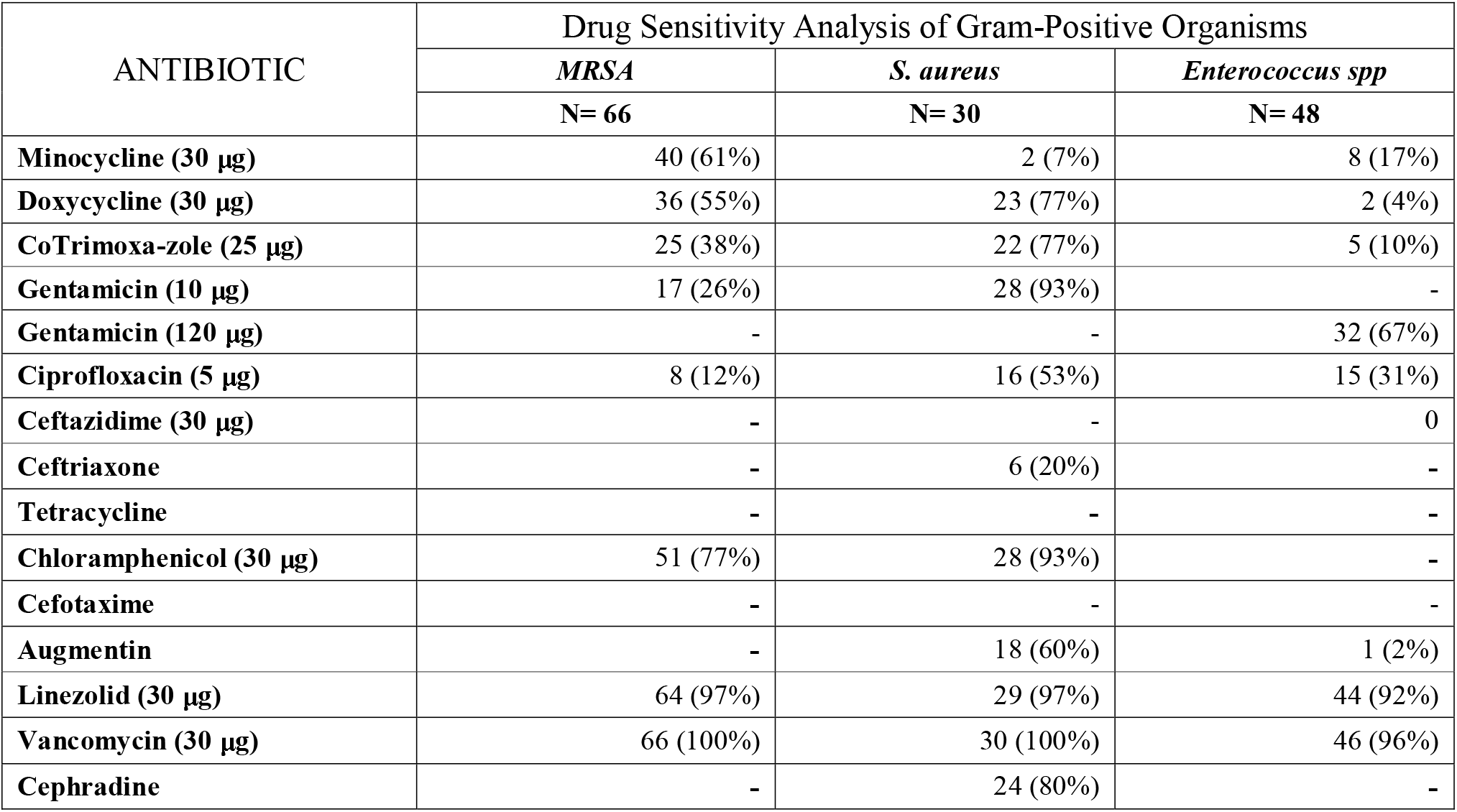
Drug Sensitivity Analysis (Gram Positive Bacteria)

According to the data from a study carried out in 2011 among Asian countries to determine the distribution of VAP and their anti-microbial resistance patterns, *Acinetobacter* spp (36.5%) was the most frequently isolated pathogen followed by *Pseudomonas aeruginosa* (25.9%) and *Klebsiella pneumoniae* (16.8%) and *Staphylococcus aureus* (12.2%). All of the isolates were resistant to important anti-microbials. A significant proportion of *Staphylococcus aureus* and *Pseudomonas aeruginosa* isolates were MDR. *Acinatobacter* spp and some *Pseudomonas aeruginosa* isolates were also resistant to Imipenem(20) A similar study conducted in the Korea in 2021 showed *Acinetobacter baumannii* as the most common pathogens isolated followed by *Staphylococcus aureus, Klebsiella pneumoniae* and *Pseudomonas aeruginosa*. Most of the isolates of *Acinatobacter baumannii* (97%) and *Staphylococcus aureus* (88%) were MDR. The pattern differed when compared to results obtained by studies conducted in the USA and UK. According to a multicenter study conducted in nine European countries, the most common pathogen isolated for nosocomial pneumonia patients were *Staphylococcus aureus* and *Pseudomonas Aeruginosa*, similar to the pattern observed in the US (22), except in Greece and Turkey where the most common pathogen was *Acinetobacter baumannii* alike our findings (23).

A few potential limitations have been identified and addressed if necessary, these may affect the interpretation and generalizability of the findings. Firstly, the study was conducted in a single tertiary care hospital in Rawalpindi, Pakistan, which may limit the results. Comparison to similar data from other hospitals or regions with different patient populations will also be necessary. These results published will serve as a point of call for other setups in the region. Secondly, the study had a limited sample size of 802 individual patient samples, which may limit the assessment on prevalence of antimicrobial resistance in the broader population. Thirdly, the study only examined tracheal tube isolates from patients with ventilator-associated pneumonia, which may not fully capture the diversity of bacterial infections in the ICU. Fourthly, the study was conducted over a period of three and a half years, which may not be long enough to fully capture changing antimicrobial resistance patterns over time.

## CONCLUSION

*Acinetobacter* spp were the most prevalent strain in the ICU tracheal tubes isolates. Increased bacterial load and resistance in the ICUs if not controlled, can lead to threatening outbreaks bringing along a disease burden our health care systems might possibly collapse under. Polymixcin B is the most effective antibiotic for use against *Acinetobacter* spp, *Pseudomonal* spp, *Escherichia coli. Pseudomonal* spp showed a high sensitivity to Tigecycline but presented a low rate of susceptibility to carbapenems, aminoglycosides and 3rd generation cephalosporins. *MRSA* showed 100% sensitivity to Vancomycin followed by Linezolid. Hence, Vancomycin and Linezolid for MRSA should depend on local resistance patterns and availability of the drugs. Malpractices in antibiotic prescription by majority of the physicians, self-medication and interrupted dosages are leading causes of heightening resistance among pathogens to antibiotics that were once considered the saving grace of mankind.

## RECOMMENDATIONS

Timely intervention in this regard via implementation of effective preventive measures and strict surveillance of ICUs, will contribute significantly in decreasing morbidity, mortality and costs of the extensive treatment that our patients and health care system have to bear. Furthermore, antibiotics are the cornerstone for treatment of infectious diseases, however the sustainability of antibiotics, a ubiquitous worldwide utility, is severely constrained due to its frequent misuse. More elaborate and long-term studies should be done to keep a check on the evolving antimicrobial sensitivity patterns for formulating better empirical therapy guidelines for local practices. Need of the moment is antimicrobial stewardship. It is pertinent that antibiotics are prescribed with responsibility, response reviewed timely, in order to reduce or replace the drug if necessary

### Limitations

Our research may have certain restrictions. Only hospital employees, retired military personnel, and their families are permitted access to this hospital. Thus, women make up a larger proportion of patients than males do. The fact that there are more ladies and children in the study than males means that it will be biased. We have two and five years’ worth of data. Because of the small sample size, there may be differences in the results.

## Data Availability

All data produced in the present study are available upon reasonable request to the authors

